# High diagnostic rate of whole genome sequencing in primary ciliary dyskinesia

**DOI:** 10.1101/2024.02.21.24302995

**Authors:** Holly A Black, Sophie Marion de Proce, Jose L Campos, Alison Meynert, Mihail Halachev, Joseph A Marsh, Robert A Hirst, Chris O’Callaghan, Scottish Genomes Partnership, Javier Santoyo-Lopez, Jennie Murray, Kenneth Macleod, Don S Urquhart, Stefan Unger, Timothy J Aitman, Pleasantine Mill

## Abstract

**Aim:** Primary ciliary dyskinesia (PCD) is a genetic disorder affecting motile cilia. Most cases are inherited recessively, due to variants in more than 50 genes that result in abnormal or absent motile cilia. This leads to chronic upper and lower airway disease, sub-fertility and laterality defects in some cases. Given overlapping clinical features and genetic heterogeneity, diagnosis can be difficult and often occurs late. Of those tested, an estimated 30% of genetically screened PCD patients still lack a molecular diagnosis. Here, we aimed to identify how readily a genetic diagnosis could be made in a clinically diagnosed population using whole genome sequencing (WGS) to facilitate identification of pathogenic variants in known genes as well as identify novel PCD candidate genes.

**Maethods:** WGS was used to screen for variants causing PCD in 8 clinically diagnosed PCD patients, sequenced as trios where parental samples were available.

**Results:** Seven of the eight cases (87.5%) had homozygous or biallelic variants in *DNAH5*, *DNAAF4* or *DNAH11* that were classified as pathogenic or likely pathogenic. Three of the variants were deletions, ranging from 3kb to 13kb, for which WGS identified precise breakpoints, permitting confirmation by Sanger sequencing. WGS yielded a high genetic diagnostic rate from this clinically diagnosed population, in part through detection of structural variants as well as identification of a *de novo* variant in a novel PCD gene *TUBB4B*.

**Conclusion:** A molecular diagnosis allows for appropriate clinical management for cases and their families, including prediction of phenotypic features correlated to genotype. Here, WGS uplifted genetic diagnosis in cases of clinically diagnosed PCD by identifying structural variants and novel modes of inheritance in new candidate genes. Our study suggests that WGS could be a powerful part of the PCD diagnostic toolkit to increase the current molecular diagnostic yield from 70%. It provides important new insight into our understanding of fundamental biology of motile cilia as well as of variation in the non-coding genome in PCD.

**Summary:** Whole genome sequencing (WGS) yielded a high genetic diagnostic rate (100%) in eight Scottish patients with clinically diagnosed primary ciliary dyskinesia (PCD) by detection of large structural variants, homology modelling and identification of a novel disease gene with a dominant mode of inheritance. Prioritised WGS may facilitate early genetic diagnosis in PCD.

## Introduction

Primary ciliary dyskinesia (PCD, OMIM: PS244400) is a genetic disorder of motile cilia (1–3), highly structurally organised organelles that project from the surface of the cells. Their organised structure allows the cilia to beat in a coordinated manner to effectively propel fluids across the surface of the airways, brain ventricles and reproductive tracts. However, in PCD, these cilia have an abnormal structure or function, resulting in static cilia, cilia that beat in an uncoordinated manner, or a complete absence of cilia (2,3). This can result in a chronic respiratory disease, due to impaired mucociliary clearance, as well as hydrocephaly, laterality defects (e.g. situs inversus) and fertility defects in a subset of patients. PCD often presents at birth as unexplained neonatal respiratory distress. Affected children then present with variable clinical features including a daily wet cough, chronic respiratory tract infections, rhinitis, sinusitis and otitis media. Over time, these can lead to debilitating long-term complications, including bronchiectasis and hearing impairment (1–6).

However, PCD is a heterogeneous disorder that has significant phenotypic overlap with other genetic respiratory diseases, such as cystic fibrosis and primary immunodeficiency. The current clinical diagnostic work-up for suspected PCD cases requires a battery of highly specialised tests (7,8). This includes detection of low nasal nitric oxide (nNO), as well as high-speed video microscopy (HSVM) to assess cilia beating pattern and frequency and transmission electron microscopy (TEM) to assess the cilia ultrastructure from a nasal brush biopsy. Abnormalities identified in cilia structure and/or beating pattern or in the presence of clinical features such as chronic oto-sinopulmonary symptoms can be used to confirm a diagnosis of PCD (1,2,6,8,9). Access to diagnostic testing is limited to a small number of highly specialised units in the UK. Families must travel for testing, often over long distances and sometimes on multiple occasions. Even in those patients with a clinical diagnosis of PCD, 21% had normal ciliary ultrastructure (10). Therefore, additional sensitive and accessible diagnostic techniques acceptable to patients of all ages are required.

PCD is a genetically heterogeneous disorder. The majority of cases are autosomal recessively inherited, with over 50 genes currently associated with PCD (1,2,6,11,12). Mutations in these genes account for approximately 70% of PCD cases screened, suggesting additional causal genes exist and/or more complicated structural variants (SVs) are being missed in known genes (13). The genes associated with PCD encode proteins involved in cilia structure, cilia assembly or regulatory complexes. There is often a correlation between the affected gene and the defects in cilia motility or ultrastructure observed at diagnosis (1,6,7,9). However, this is not always the case: it is estimated that 9-20% of PCD cases have normal or inconclusive ultrastructure by TEM (14). Similarly normal or non-diagnostic HSVM results complicate diagnosis for a significant portion of known PCD genes (15). nNO is also considered a useful screening test prior to referral for nasal brushing. While low nNO measurements may be indicative of PCD in patients lacking clear ultrastructural defects, its use, particularly in young children, has not been universally rolled out across PCD centres (16,17) and patients with mutations in at least some PCD genes show normal nNO levels (18,19). In such suspected cases, where a clinical diagnosis is unclear, a genetic diagnosis is an important means to confirm PCD (1,5,6,20). However, genetic testing often occurs late in the diagnostic work-up for PCD (7). Recent guidelines have recommended studies to investigate the utility of genetic testing in the PCD diagnostic pathway, particularly as next generation sequencing (NGS) is increasingly available (8,21,22).

In this study, we investigated the utility of whole genome sequencing (WGS) to detect disease-causing variants in children and young adults with a clinical diagnosis of PCD. We selected a non-endogamous population where PCD was clinically confirmed to test whether WGS could effectively improve the molecular diagnostic yield of PCD. WGS would provide unbiased genome coverage beyond exons of ∼50 genes associated with PCD whilst simultaneously detecting a range of variants, from single nucleotide changes, small indels as well as complicated SVs, which may be missed by targeted sequencing approaches. Eight patients were recruited, alongside parental samples where available. This led to a genetic diagnosis for all eight cases, three of whom had a pathogenic deletion of 3 kb-13 kb and one patient had a de novo missense variant p.P259L in a novel candidate gene *TUBB4B*.

## Materials and Methods

### Patient cohort

Eight patients with a confirmed clinical diagnosis of PCD (50% female), aged 6 to 31 years, were recruited to the study. Signed and informed consent was obtained from the affected individual as well as relatives through approved protocols. Sample IDs were assigned and the key known only those within the research group. Clinical phenotypes are shown in **Table 1**. For details of blood samples, DNA extraction, and analysis of sample ethnicity see Supplementary Methods and **Supplementary Figure 1**. The study was approved by the London-West London and Gene Therapy Advisory Committee Research Ethics Committee (REC number 11/LO/0883).

**Table 1:**
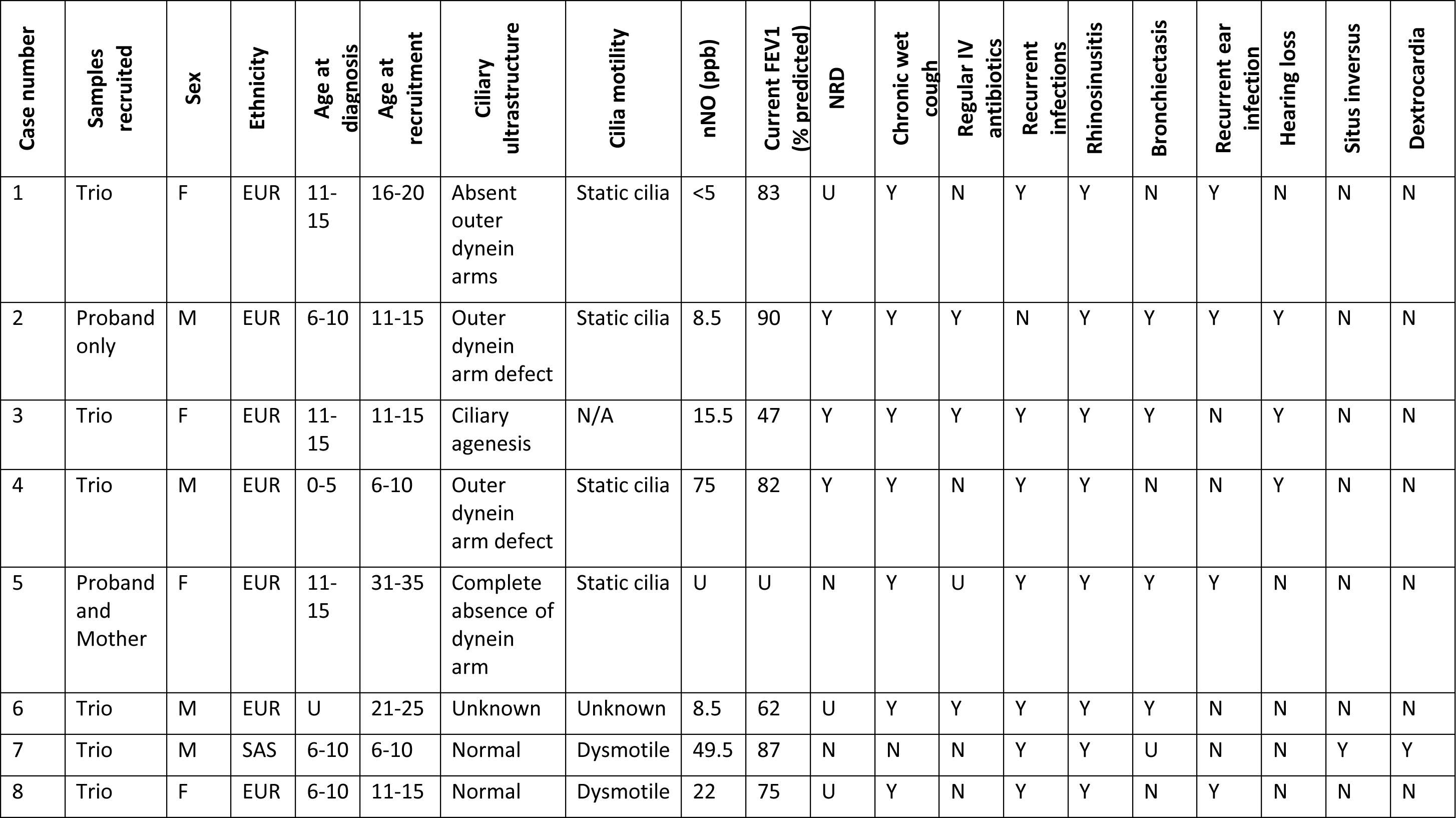
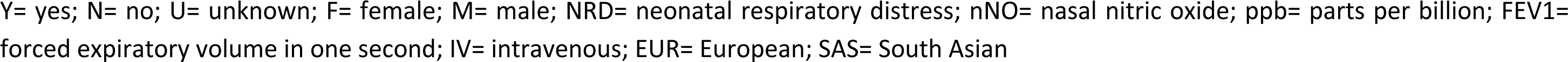
Clinical details of PCD cases.

| Case number | Samples recruited | Sex | Ethnicity | Age at diagnosis | Age at recruitment | Ciliary ultrastructure | Cilia motility | nNO (ppb) | Current FEV1 (% predicted) | NRD | Chronic wet cough | Regular IV antibiotics | Recurrent infections | Rhinosinusitis | Bronchiectasis | Recurrent ear infection | Hearing loss | Situs inversus | Dextrocardia |
| --- | --- | --- | --- | --- | --- | --- | --- | --- | --- | --- | --- | --- | --- | --- | --- | --- | --- | --- | --- |
| 1 | Trio | F | EUR | 11-15 | 16-20 | Absent outer dynein arms | Static cilia | <5 | 83 | U | Y | N | Y | Y | N | Y | N | N | N |
| 2 | Proband only | M | EUR | 6-10 | 11-15 | Outer dynein arm defect | Static cilia | 8.5 | 90 | Y | Y | Y | N | Y | Y | Y | Y | N | N |
| 3 | Trio | F | EUR | 11-15 | 11-15 | Ciliary agenesis | N/A | 15.5 | 47 | Y | Y | Y | Y | Y | Y | N | Y | N | N |
| 4 | Trio | M | EUR | 0-5 | 6-10 | Outer dynein arm defect | Static cilia | 75 | 82 | Y | Y | N | Y | Y | N | N | Y | N | N |
| 5 | Proband and Mother | F | EUR | 11-15 | 31-35 | Complete absence of dynein arm | Static cilia | U | U | N | Y | U | Y | Y | Y | Y | N | N | N |
| 6 | Trio | M | EUR | U | 21-25 | Unknown | Unknown | 8.5 | 62 | U | Y | Y | Y | Y | Y | N | N | N | N |
| 7 | Trio | M | SAS | 6-10 | 6-10 | Normal | Dysmotile | 49.5 | 87 | N | N | N | Y | Y | U | N | N | Y | Y |
| 8 | Trio | F | EUR | 6-10 | 11-15 | Normal | Dysmotile | 22 | 75 | U | Y | N | Y | Y | N | Y | N | N | N |

Y= yes; N= no; U= unknown; F= female; M= male; NRD= neonatal respiratory distress; nNO= nasal nitric oxide; ppb= parts per billion; FEV1= forced expiratory volume in one second; IV= intravenous; EUR= European; SAS= South Asian

### Whole Genome Sequencing

DNA was sequenced by WGS at Edinburgh Genomics. Libraries were prepared using the Illumina TruSeq PCR-free protocol and sequenced on the Illumina HiSeq X platform. The average yield per sample was 136 Gb, with mean coverage of 36x (range 33.9-38.3).

### Gene Panel, data analysis and variant classification

A virtual gene panel of 146 genes was created, which included the known ‘green’ 34 PCD genes plus 107 suspected ciliopathy genes that may have respiratory features based on the PCD PanelApp panel (v1.14) with five additional genes identified in the literature (*CFAP300*, *DNAH6*, *DNAJB13*, *STK36* and *TTC25)* (23–25). Analysis of variants was carried out as described in Supplementary Methods. Additional analyses were carried out across the *FOXJ1* locus for Case 3, in whom no diagnostic variants were identified using the virtual gene panel. A genome-wide expanded analysis identified a *de novo* missense mutation p.P259L (chr9:g.137242994:C>T (hg38)) in the gene *TUBB4B* only in the patient, and not present in either parent (26).

### Modelling of whole exome sequencing data

A whole exome sequencing (WES)-like subset of the WGS data was obtained by extracting only the reads mapping to the regions in the TWIST Exome Capture Kit (using samtools v1.6) from the BAM file for each sample. WES CNV calling was performed by ExomeDepth (v1.1.15) as detailed in Supplementary Methods.

### Homology modelling of DNAH11 and location of missense variants

A homology model of the C-terminal region of the DNAH11 motor domain (residues 3348-4504) was built using PHYRE2 (27,28). The effects of mutations in the C-terminal domain (CTD) (residues 4124-4504) on protein stability were modelled with FoldX (29). Sequences of human dynein genes were aligned with MUSCLE (30) and the sequence alignment was visualised with MView (31). Further details are given in the Supplementary Methods.

## Results

Eight PCD patients were recruited to the study as six trios, one proband-mother duo and one singleton (**Table 1**). All eight cases had a confirmed clinical diagnosis of PCD, following a nasal brush biopsy. In all cases, nasal brushings were undertaken for clinical suspicion of PCD. Presentations and phenotype severity were varied and are summarised in **Table 1**.

Using a virtual PCD panel approach to initially screen the WGS, we confirmed genetic diagnosis for seven of the eight cases (**Table 2**). Three cases had biallelic pathogenic and/or likely pathogenic variants in the outer dynein arm heavy chain subunit gene *DNAH5*, which has previously been shown to contribute to the largest proportion of PCD cases among populations of European descent (1). Case 1 had a hemizygous nonsense variant (c.5281C>T, p.(Arg1761Ter)), inherited from the mother and previously reported in PCD cases (32,33), and an overlapping 13kb deletion on the other allele, inherited from the father (**Supplementary Figure 3**). Case 2 had two nonsense variants in *DNAH5*, c.3949C>T, p.(Gln1317Ter), which was previously reported (34), and the c.5281C>T, p.(Arg1761Ter) variant. ddPCR was used to phase the variants in Case 2 (**Supplementary Figure 4**). Case 4 had a single base pair deletion, resulting in a frameshift and premature termination codon (PTC) (c.10815del, p.(Pro3606Hisfs)), and a single base pair duplication, resulting in direct creation of a PTC (c.13458dup, p.(Asn4487Ter)), which were shown to be in trans through trio-based phasing. Both variants have been reported previously in PCD cases (33). All three cases with biallelic variants in *DNAH5* were shown to have absence or defects of outer dynein arms on TEM (**Figure 1**), with Case 1 and Case 2 also shown to have static cilia (**Table 1, Supplementary Video 1**). This is consistent with loss of DNAH5, which has been shown to result in outer dynein arm truncation and immotile cilia (33). All three share a similar clinical phenotype, with chronic wet cough, sinus problems and recurrent chest infections. Hearing loss was also noted in cases 2 and 4.

**Table 2:** Genetic variants identified in PCD cases. Variants were classified according to the ACMG/ACGS guidelines (38,39); the criteria met for each variant are shown.

| Case number | Gene | Transcript | dbSNP ID | Variant(s) | Zygosity | gnomAD allele frequency (v4.0.0) | In silico predictions | ACMG/ACGS Classification | ClinVar Accession Number |
| --- | --- | --- | --- | --- | --- | --- | --- | --- | --- |
| 1 | DNAH5 | NM_001369.3 | N/A | c.5272-955_6197del p.(?) | Compound heterozygous | Absent<sup>a</sup> | N/A | P (PVS1, PM3, PP4) | SCV001334255 |
|  |  |  | rs148891849 | c.5281C>T, p.(Arg1761Ter) (32-34) |  | 6x10<sup>-5</sup> | N/A | P (PVS1, PM2, PM3_str, PP4) | SCV001334256 |
| 2 | DNAH5 | NM_001369.3 | rs1769508571 | c.3949C>T, p.(Gln1317Ter)(74) | Compound heterozygous | 1.6x10<sup>-6</sup> | N/A | P (PVS1, PM2, PM3_str, PP4) | SCV001334257 |
|  |  |  | rs148891849 | c.5281C>T, p.(Arg1761Ter)(32-34) |  | 6x10<sup>-5</sup> | N/A | P (PVS1, PM2, PM3_str, PP4) | SCV001334256 |
| 3 | TUBB4B | NM_006088.6 | N/A | c.776C>T, p.(Pro259Leu) (26) | Heterozygous | Absent | Grantham Score 98 AlphaMissense 0.998 (LP) | P (PS2, PM2, PP2, PP3) | SCV002770069.1 |
| 4 | DNAH5 | NM_001369.3 | rs397515540 | c.10815del, p.(Pro3606Hisfs) (33) | Compound heterozygous | 4.1x10<sup>-4</sup> | N/A | P (PVS1, PM2, PM3_str, PP4) | SCV001334258 |
|  |  |  | rs775696136 | c.13458dup, p.(Asn4487Ter) (33) |  | 1.6x10<sup>-4</sup> | N/A | P (PVS1, PM2, PM3_str, PP4) | SCV001334259 |
| 5 | *DNAAF4* | NM_130810.4 | N/A | c.784-1037_894-2012del p.(?) (35) | Homozygous | $2.5 \times 10^{-4b}$ | N/A | P (PVS1, PM3_str, PP4) | SCV001334260 |
| 6 | *DNAAF4* | NM_130810.4 | rs770136467 | c.856G>T, p.(Glu286Ter) | Compound heterozygous | $2.6 \times 10^{-5}$ | N/A | P (PVS1, PM2, PP4) | SCV001334261 |
|  |  |  | N/A | c.1048-149_*1048del p.(?) |  | Absent<sup>a</sup> | N/A | P (PVS1, PM3, PP4) | SCV001334262 |
| 7 | *DNAH11* | NM_001277115.2 | rs72658835 | c.13373C>T, p.(Pro4458Leu) (36,37,47,51) | Homozygous | $8.4 \times 10^{-5}$ | Grantham Score 98<br>REVEL 0.377 (Uncertain)<br>AlphaMissense 0.625 (LP) | LP (PM2, PM3, PP3, PP4) | SCV001334263 |
| 8 | *DNAH11* | NM_001277115.2 | rs1450540788 | c.10221_10222del, p.(Cys3409Trpfs) | Compound heterozygous | $1.5 \times 10^{-5}$ | N/A | P (PVS1, PM2, PP4) | SCV001334264 |
| | | | N/A | c.13288G>C, p.(Gly4430Arg) (75) | | $2 \times 10^{-6}$ | Grantham score 125<br>REVEL 0.641 (Damaging)<br>AlphaMissense 0.813 (LP) | LP (PM1_sup, PM2, PM3_str, PP3, PP4) | SCV001334265 |

P= pathogenic; LP= likely pathogenic; N/A= not applicable. <sup>a</sup>absent from gnomAD SV and gnomAD CNV. <sup>b</sup>frequency from gnomAD SV.

**Figure 1:**
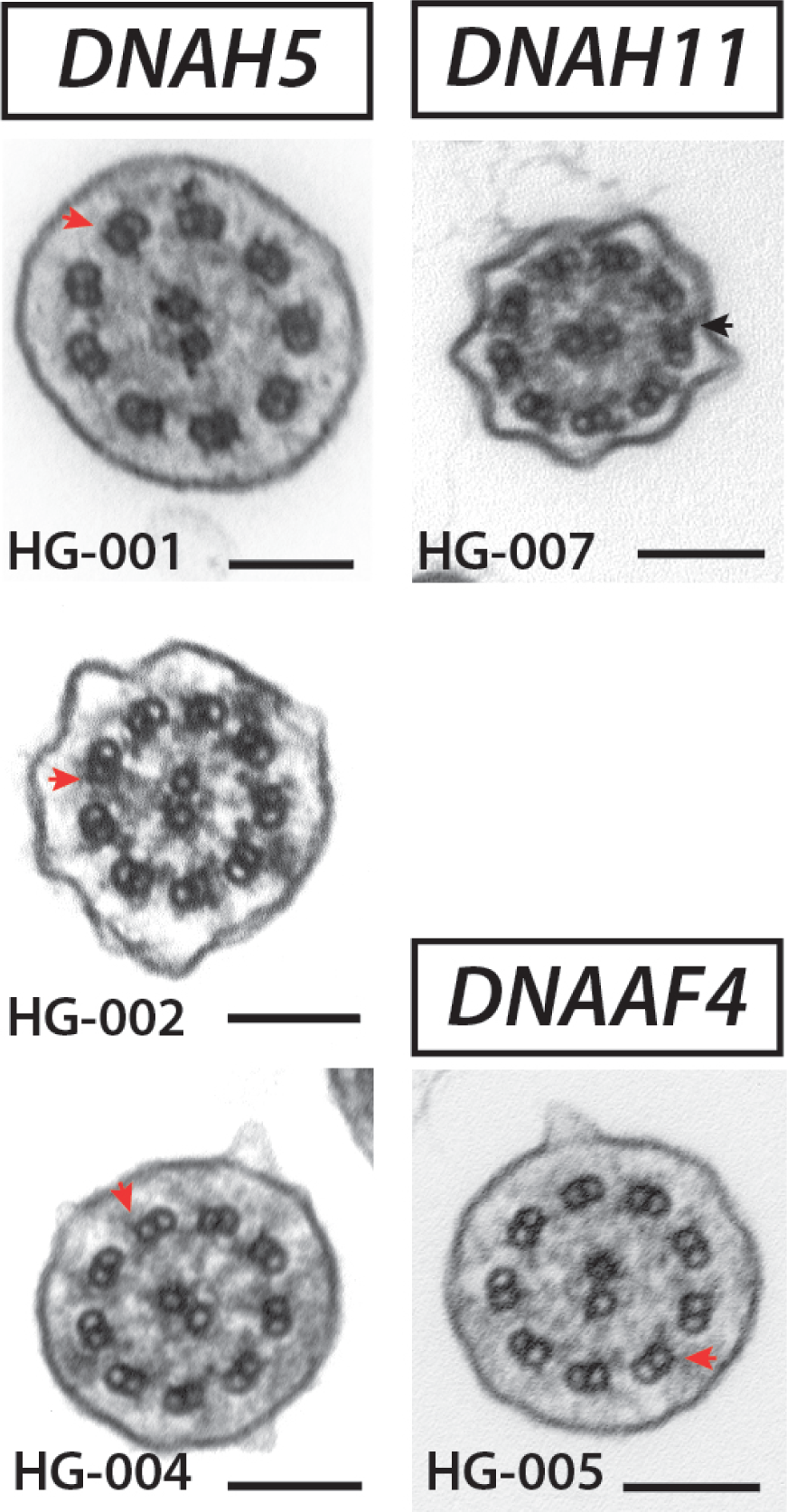
Ultrastructure analysis supports genetic diagnosis for outer arm dynein variants in PCD. (left) *DNAH5* variants disrupt outer dynein arms (red arrowhead = disrupted, black arrowheads normal) across axonemes of nasal brush samples from cases HG-001, HG-002 and HG-004. (right) In contrast, *DNAH11* variants do not uniformly disrupt outer dynein arms (black arrowheads) in cilia from nasal brush of case HG-007. Disruption of both inner and outer dynein arms (red arrowheads) is observed in *DNAAF4* variants, as shown for HG-005. Scale bars = 100 nm.

Two cases had likely pathogenic variants in the cytoplasmic axonemal dynein assembly factor *DNAAF4* (*DYX1C1*), necessary for assembly and/or stability of the inner and outer dynein arms (**Table 2**) (35). Case 5 had a homozygous 3.5kb deletion encompassing exon 7 of *DNAAF4*, which was also heterozygous in the mother. There was no evidence of uniparental disomy and, given Case 5 is also reported to have an affected sibling, it is likely the father also carries the 3.5kb deletion, although the father’s DNA was unavailable. This variant has previously been reported in PCD (35). Ultrastructural analysis was consistent with previous reports for *DNAAF4* mutations, with reports of absent dynein arms (35) (**Figure 1**) and immotile cilia shown on HSVM. Case 6 had a maternally-inherited nonsense variant (c.856G>T, p.(Glu286Ter)) and a paternally-inherited 3.1 kb deletion encompassing the last two exons of *DNAAF4*, predicted to disrupt the C-terminal tetratricopeptide-like helical domain region (TPR) by deleting the final 71 amino acids, as well as the 3’UTR. We predict this allele to be loss-of-function as the resulting transcript is predicted to be subject to nonsense-mediated decay. Case 6 has bronchiectasis with a recurrent need for antibiotics. He has no ear or hearing problems and situs solitus.

A further two cases had pathogenic or likely pathogenic variants in the outer dynein arm heavy chain subunit *DNAH11*. Case 7 had a homozygous missense variant in *DNAH11* (c.13373 C>T, p.(Pro4458Leu)) previously reported in PCD cases (36,37), at a low frequency in gnomAD and *in silico* predictions support pathogenicity (**Table 2**). There was sufficient evidence to classify the variant as likely pathogenic using ACMG guidelines (38,39). Case 8 had a heterozygous 2 bp deletion, resulting in a PTC (c.10221_10222del, p.(Cys3409Trpfs)), which was classified as pathogenic (**Table 2**). The second variant was a heterozygous missense (c.13288G>C, p.(Gly4430Arg)). This variant is at a low frequency in gnomAD and *in silico* predictions support pathogenicity, but it has not previously been reported in PCD cases. As the variant is in trans with the c.10221_10222del variant, shown by phasing with parental samples, there is sufficient evidence to classify the variant as likely pathogenic (**Table 2**). Both Case 7 and Case 8 were shown to have a normal cilia ultrastructure with a dysmotile phenotype (**Figure 1, Supplementary Video 2**), consistent with loss of DNAH11, which localises to the proximal portion of respiratory cilia (40). There is however phenotypic variability between Case 7 (situs inversus with dextrocardia and recurrent chest infections) and Case 8 (chronic wet cough and recurrent ear infections, with situs solitus).

The two missense variants in *DNAH11* both reside at the 3’ C-terminal domain (CTD) of the axonemal dynein heavy chain. While there is no structure available for DNAH11, there are homologous structures of other dynein proteins. To investigate the effects of the *DNAH11* missense mutations on protein structure, we built a homology model of the DNAH11 motor domain. Both mutations occur within the CTD, on the outer side of the dynein motor dimer (**Figure 2A**) and are intriguingly very close to each other in three-dimensional space (**Figure 2B**), separated by only 4.2 Å.

**Figure 2:**
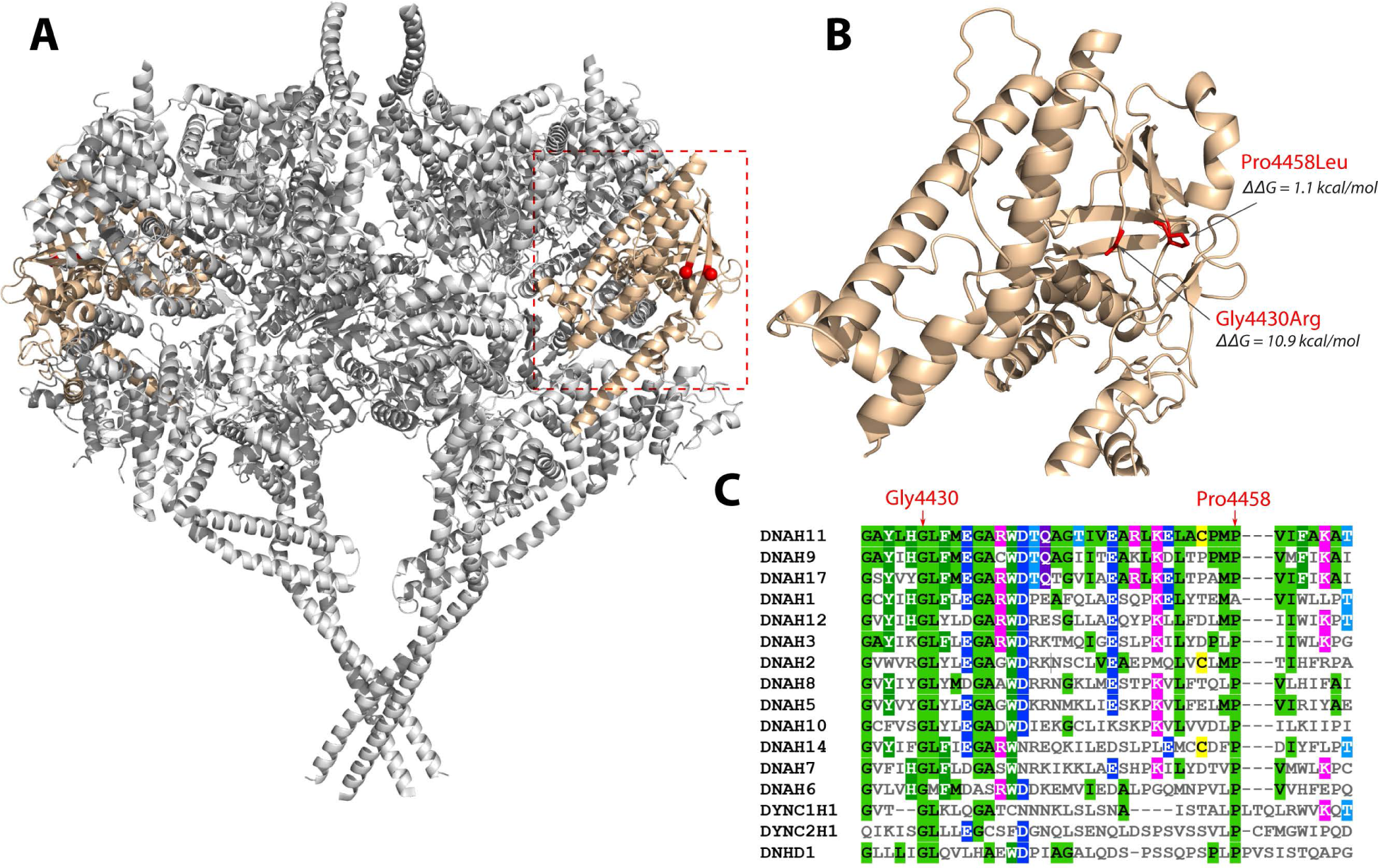
Structural and evolutionary analysis of DNAH11 missense mutations. (A) Structure of the human cytoplasmic dynein-1 dimer (PDB ID: 5NUG), with the location of the C-terminal domain coloured beige, and the equivalent sites of the DNAH11 mutations highlighted in red. (B) Homology model of the DNAH11 C-terminal domain with the sites of the missense mutations shown in red, along with the ΔΔG values calculated with FoldX. (C) Multiple sequence alignment of human dynein proteins around the region where the missense mutations occur.

Molecular modelling of the missense mutations using the program FoldX (29) predicts that the Gly4430Arg should be extremely disruptive to protein structure, with a ΔΔG of 10.9 kcal/mol. We also modelled all other CTD missense variants presented in the gnomAD v2.1 database (41). Remarkably, out of 269 variants (**Supplementary Table 1**), Gly4430Arg has the highest ΔΔG and therefore is predicted to be the most damaging. Moreover, this position is fully conserved across all human dyneins (**Figure 2C**). This strongly suggests that Gly4430Arg is pathogenic due to its disruptive effects on protein structure.

In contrast to Gly4430Arg, Pro4458Leu is predicted to be relatively mild at a structural level, with a ΔΔG of 1.1 kcal/mol, making it only the 65^th^ most damaging out of 269 variants (**Supplementary Table 1**). However, this residue is highly conserved, existing as a proline across all human dyneins except DNAH1, where it is an alanine. Thus, while Pro4458 is unlikely to cause a severe destabilisation of protein structure, its remarkable proximity to Gly4430 combined with a moderate structural perturbation and high conservation are supportive of pathogenicity.

To assess whether the three deletion variants identified in Cases 1, 5 and 6 would have been identified using WES, as opposed to WGS, the WGS data was subsetted to create a WES-like dataset for each sample (**Supplementary Figure 2**). CNV calling using ExomeDepth was able to identify the *DNAH5* variant c.5272-955_6197del p.(?) in both the proband and father for Family 1. However, the deletion, which spans five exons and has a breakpoint within exon 37, was called as covering only 4 of the 5 affected exons in the proband. The single exon deletion of *DNAAF4* c.784-1037_894-2012del p.(?) was identified for the proband in Family 5, where it is homozygous, but not in the heterozygous mother. The *DNAAF4* deletion c.1048-149_*1048del p.(?) identified in Family 6 was not detected in the proband or father, despite both carrying the variant.

The phenotype of Case 3 is particularly relevant as ultrastructural analysis revealed ciliary agenesis (**Supplementary Video 3**), sometimes referred to as reduced generation of multiple motile cilia (RGMC), a specific subtype of PCD. In addition, Case 3 has shunted hydrocephalus, having undergone initial ventriculo-peritoneal (VP) shunt insertion neonatally, and a subsequent VP shunt revision as an adolescent. To date, few genes have been implicated in this RGMC phenotype by recessive inheritance, *CCNO* (cyclin O) and *MCIDAS* (multicilin) (42,43) but no pathogenic or potentially pathogenic variants were identified in either gene. Heterozygous de novo mutations in the master motile ciliogenesis transcriptional regulator *FOXJ1* were identified as the first autosomal dominant cause of a distinct PCD-like condition, associated with chronic respiratory disease, laterality defects and hydrocephalus (18). Similar cellular defects are observed with reduced apical docking of centrioles and fewer cilia, but a focused analysis revealed no identifiable pathogenic mutations in the *FOXJ1* locus. An expanded, whole genome analysis for SNP and indel candidates produced a very limited list of variants (**Supplementary Table 2**). This included a de novo missense mutation p.P259L (chr9:g.137242994:C>T (hg38)) in the gene *TUBB4B* only in the patient, and not present in either parent or found on gnomAD 4.0 or other publicly available databases. Whilst interpretation of pathogenicity from a single patient is limiting, as part of a large international collaboration, we were able to identify a further eleven patients with PCD carrying *TUBB4B* variants identified by next-generation sequencing (NGS) (26). This included five patients with PCD-only carrying the identical p.P259L (chr9:g.137242994:C>T) variant, one carrying a different missense p.P259S (chr9:g.137242993:C>T (hg38)) variant and one patient carried an in-frame ten amino acid duplication p.F242_R251dup (chr9: g.137242941_137242970dup (hg38)) (26). Moreover, we also identified a recurrent de novo *TUBB4B* variant four patients with a p.P358S (chr9:g.137243290:C>T (hg38)) variant, who presented with features of both PCD with Leber congenital amaurosis and sensorineural hearing loss. In-depth functional analyses in cell and animal models confirmed pathogenicity and distinct dominant-negative mechanisms of action to cause different disease presentations (26). In summary, our WGS strategy was very effective at identifying a novel PCD disease gene with the first report of dominant negative disease mechanisms.

## Discussion

In this study, WGS of affected probands and family members led to a genetic diagnosis in all eight PCD patients, representing 11 different mutations in 3 autosomal recessive PCD genes and 1 de novo mutation in a novel autosomal dominant PCD candidate *TUBB4B*. In Case 3, a patient with features of reduced generation of motile cilia and hydrocephalus, no pathogenic variants would been identified in a panel-based approach, even with targeted sequencing of potential candidates for known RGMC loci. The diagnostic rate of 100% in our small study is higher than previous reports, in which 60-70% of cases received a genetic diagnosis based on the known PCD gene panels (1,5,12,13,20), 75% on extended NGS panels (44) and 68-94% by WES (45–48).

Next-generation sequencing (NGS) technologies continue to revolutionise rare genetics research and clinical diagnostics, where the advantages of WES versus WGS are often fiercely debated. Cheaper in terms of costs of sequencing, analysis and data storage, WES is generally preferred as a front-line diagnostic tool. WES, however, has several issues in terms of evenness of genome coverage and sequence bias, particular for copy number variations (CNV). In comparison, several studies have found more accurate variant calls as well as even and unbiased coverage of coding regions are generated by WGS (49,50). Our analysis was focused on known and candidate PCD genes, screening simultaneously for single nucleotide variants (SNVs), small indels and more complex SVs. In three cases, pathogenic deletions were identified, ranging in size from 3kb to 13kb. Modelling of our WGS data to represent WES-like data suggested that WES data would only have identified the deletion in one of these three cases, in which the deletion was homozygous. Since our WES-like model has more uniform coverage than true WES data, our WES-like model could be considered more reliable for CNV calling than real-life WES sequence. However, we acknowledge that our model is based on 36X WGS data, whereas WES would be nearer 100X in practice but coverage does vary across commercial platforms. The ability to detect SVs in PCD is of importance and consistent with recent diagnostic guidelines for PCD, which highlighted that causal SVs and intronic mutations can be missed due to the large number and size of PCD genes (8,21). Further, WES would not have provided the precise breakpoint information that allowed confirmation of the three deletion variants by Sanger sequencing.

Where WGS was clearly advantageous was PCD disease gene discovery in patients without biallelic variants in known genes or in the one recent example of autosomal dominant inheritance (*FOXJ1*) or in the few cases of X-linked recessive inheritance (*RPGR, PIH1D3, OFD1)* (*12*). In Case 3, WES would have likely identified the de novo SNV in *TUBB4B,* as WES panels identified SNVs in the other 11 *TUBB4B* patients (26). Here, the clear advantage of WGS was to rule out potential non-coding alterations in the known RGMC PCD genes of known inheritance such as *FOXJ1* modes as to quickly prioritise novel candidates in our first proband. As such, a clear benefit of WGS, similar to WES, is that its findings are future-proof; the data generated can be re-screened for PCD-causing variants identified subsequent to genetic testing, if initial testing is inconclusive. However, only WGS will allow future analysis of non-coding genome for variations in regulatory elements such as of transcription factor binding sites that may underlie a subset of unsolved PCD cases.

Whatever the modality, increased genetic testing for PCD is critical. Currently genetic testing for PCD sits as an additional step to confirm diagnosis by both European and American guidelines (8,21,22). As shown here, and elsewhere, genetic testing can accurately diagnose PCD where standard clinical procedures are unavailable, impractical or inconclusive (45–47,51) Importantly, a delayed PCD diagnosis is associated with worse prognosis (52,53). A clinical diagnosis of PCD requires specialised, invasive testing, and frequently necessitates travel over considerable distances to specialist centres. These tests are unsuitable for critically ill neonates and for those unable to travel or unwilling to undergo invasive testing. Genetic testing for PCD may therefore have utility in the neonatal period or in infancy that may have been preferred to the diagnostic odyssey of the patients in this study, where the age of diagnosis was between 5 and 12 years. Suggested clinical criteria for genetic testing in PCD could include term babies who become unwell at >12 hours of age with respiratory disease, lobar collapse, situs inversus and/or an unexpectedly high or prolonged oxygen requirement (54,55). Increasing availability, reducing costs and recognised clinical utility of NGS platforms such as WGS in the neonatal and paediatric intensive care unit setting would be consistent with such indications (50,51,54,56–59). Moreover, either WES or WGS would help rule out other confounding clinical presentations such as primary immune deficiency disorders (PIDs), with overlapping symptoms of frequent, often severe, airway infections as well as recurrent otitis media, and sinusitis (60–62). Both platforms are advantageous as they allow analysis of all potential causative genes, known and novel, thus faster to keep up with recently reported genes that may not yet be included on PCD-gene panels. Indeed such technologies may prove to be more cost- and time-effective as providing a molecular diagnosis test for PCD than conventional clinical gene panels (45,46,48,63). A prospective study would appear to be warranted, to define the optimum criteria for genetic testing and diagnostic yield in neonates and older infants and children with respiratory symptomatology to help expedite PCD diagnosis.

While earlier genetic testing for PCD is clearly a priority, PCD also remains underdiagnosed. PCD has an estimated incidence of 1 in 7,500 births, rising to 1 in 2,300 in endogamous populations (64,65). In North America, it is estimated that only 1,000 patients have a confirmed diagnosis of PCD as opposed to the predicted ∼25,000-50,000 these rates would suggest to be affected with PCD (66). Similar underdiagnosis is reported in the UK, where these prevalence estimates suggest there should be at least 8,900 people with PCD in the UK; less than a quarter of these are known to the NHS highly specialised PCD service (67). A large portion of these missing patients are likely adult patients within primary care or bronchiectasis clinics, as suggested by one study which found 12% of bronchiectasis patients had pathogenic variants in known motile ciliopathy genes (68). Increased genetic testing as a first-pass diagnostic test in these suspected cases of PCD could be a way of controlling access to more labour-intensive clinical and pathological diagnostic work-up in limited specialist centres.

PCD needs to enter the precision medicine era. A genetic diagnosis is key to improved patient prognosis as we better understand genotype-phenotype relations (69) and critically to being trial-ready as much-needed genetic therapies come online (70,71). Increased genetic testing is also being combined with a curated worldwide database, similar to that of CFTR2 for Cystic Fibrosis (72), to enable a better understanding of these genotype-phenotype relations in PCD called CiliaVar (73). Such a database will significantly facilitate interpretation of variants, of which 21% has been suggested to be variants of unknown significance (VUS) (73). For example, in Cases 7 and 8 in our study, structural modelling of the CTD of *DNAH11* aided the assignment of pathogenicity of two missense variants, with one predicted to be detrimental to protein stability and the second, in close proximity to the first, shown to be highly conserved.

In conclusion, this study demonstrates the benefits of using WGS to obtain a genetic diagnosis for PCD, through its ability to detect SNVs and SVs simultaneously as well as detecting variants in genes outwith current gene panels. The detection of multi-kilobase deletions in three of the seven diagnosed cases highlights the need to detect SVs as part of the genetic testing for PCD. It also allowed rapid prioritisation of SNVs in novel candidate genes with dominant modes of inheritance. Practically and financially, WGS would likely sit behind current standard of care panel-based or WES diagnostic platforms. Given the high genetic diagnostic rate observed here and elsewhere (44,45,47,48), we suggest that genetic testing should be an early step in the current diagnostic pathway for PCD, particularly in cases where nasal brush biopsy is unavailable. By moving clinical and genetic diagnostic pathways in PCD earlier, ideally to early life, we could have a transformative long-term reduction in morbidity with access to specialist care and disease-modifying therapies commenced before permanent lung damage.

## Data Availability

All data produced in the present study are available upon reasonable request to the authors.

## Acknowledgements

We thank the PCD families who participated in this study and the UK PCD Family Support Group for support; Dr Lee Murphy and colleagues at the Wellcome Trust Clinical Research Facility; Edinburgh Genomics for sequencing and analysis; and the Brompton Hospital PCD Diagnostic Service and South East Scotland Genetics Service for clinical support.

## Supplementary Material and Methods

### SUPPLEMENTARY MATERIALS

**Supplementary Table 1: Ranked list of most disruptive reported variants (ΔΔG) in the C-terminal domain (CTD) of DNAH11 as predicted by FoldX.** ΔΔG represents the change in free energy by mutation/design of proteins as predicted by the FoldX algorithm, where ΔΔG = ΔGfold(mutation) − ΔGfold(wild type).

**Supplementary Table 2: Genome wide list of variants detected in patient HG-003 by Slivar.**

**Supplementary File 1: Gene panel used for variant filtering with G2P under a biallelic inheritance model**

**Supplementary File 2: Gene panel used for variant filtering with G2P under a monoallelic inheritance model**

**Supplementary File 3 Droplet digital PCR for variant phasing.**

**Supplementary Video 1: HSVM for cases 1, 2 and 4, which have a genetic diagnosis in *DNAH5*, showing static cilia**

**Supplementary Video 2: HSVM for cases 7 and 8, which have a genetic diagnosis in *DNAH11*, showing dysmotile cilia**

**Supplementary Video 3: HSVM for case 3, showing ciliary agenesis**

**Supplementary Figure 1:**
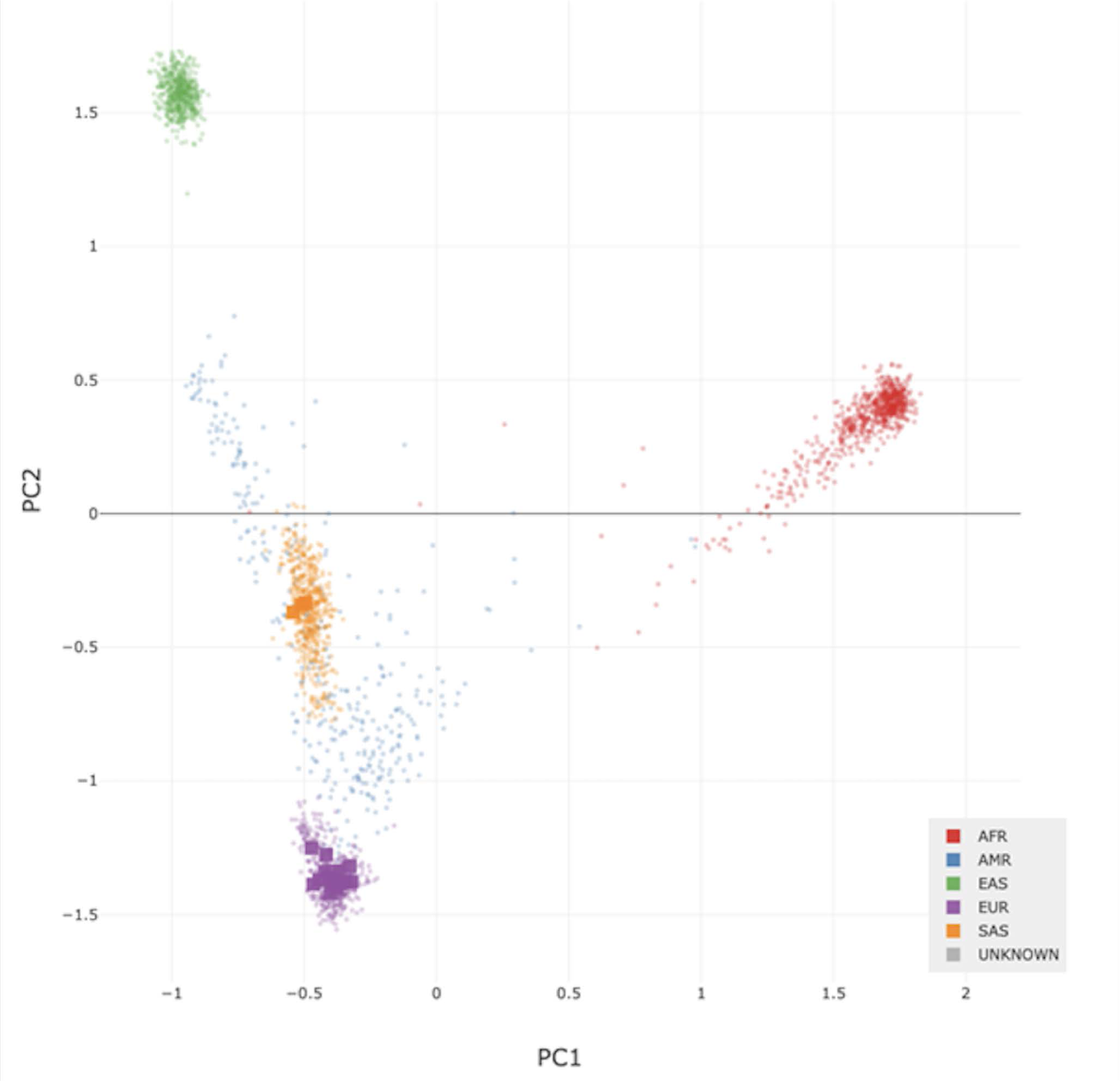
Principal component analysis (PCA) of study samples compared to 1000 Genomes project samples. Peddy was used to predict ancestry of the samples used in the study by comparison with the 1000 Genomes samples. The 1000 Genomes samples (dots) are colour-coded by location. The samples in our study are represented by squares. All samples were predicted to be of European ancestry (purple squares), except three (orange squares), which were predicted to have South Asian ancestry. AFR= African; AMR= American; EAS= East Asian; EUR= European; SAS= South Asian.

**Supplementary Figure 2:**
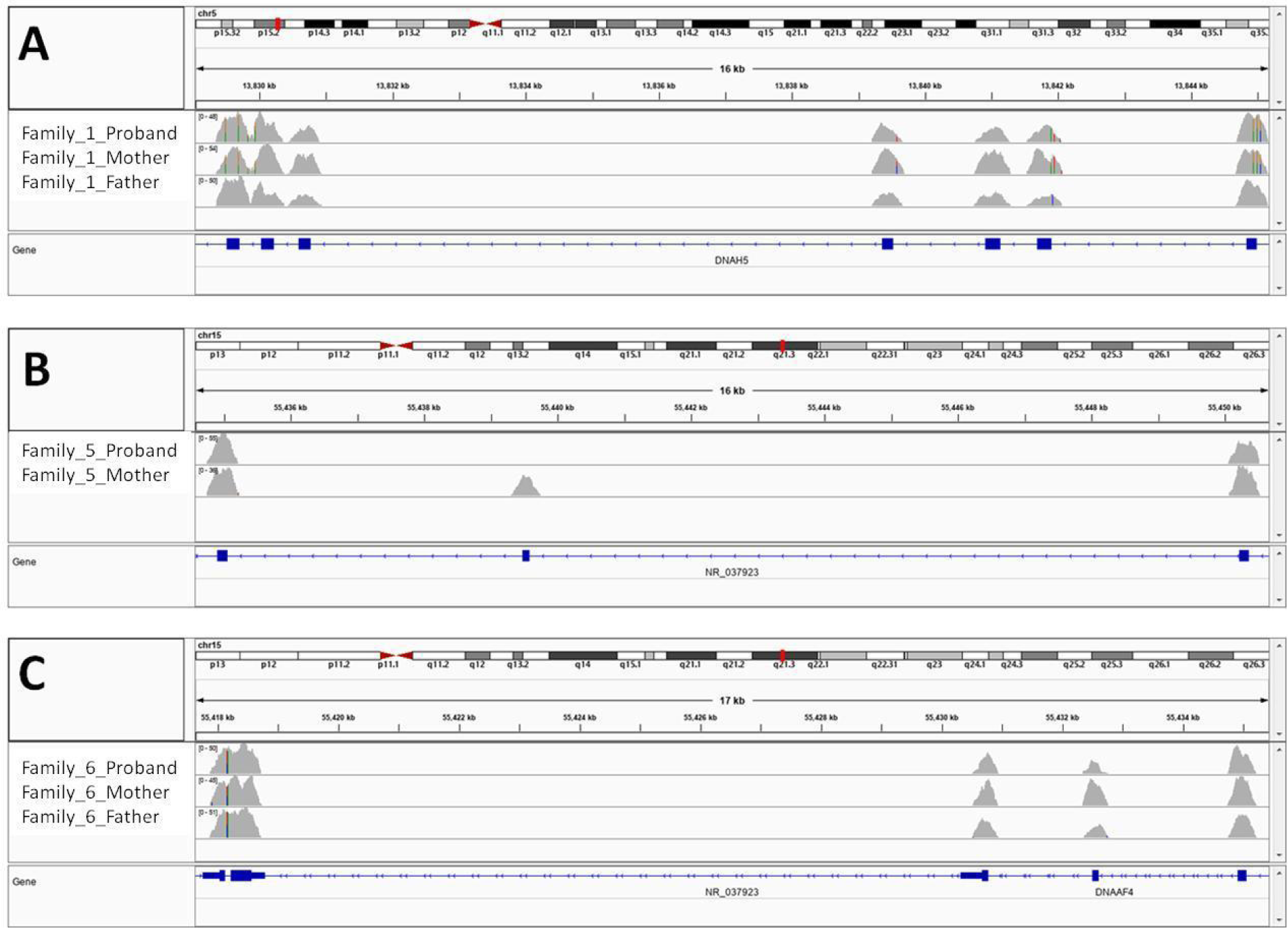
Alignments of the modelled WES data showing sufficient coverage to call the deletion variants identified in Cases 1, 5 and 6. A: Alignment for Family 1, showing coverage of exons 32-38 of *DNAH5.* B: Alignment for Family 5, showing coverage of exons 6 to 8 of *DNAAF4.* C: Alignment for Family 6, showing coverage exons 8-10 of *DNAAF4*.

**Supplementary Figure 3:**
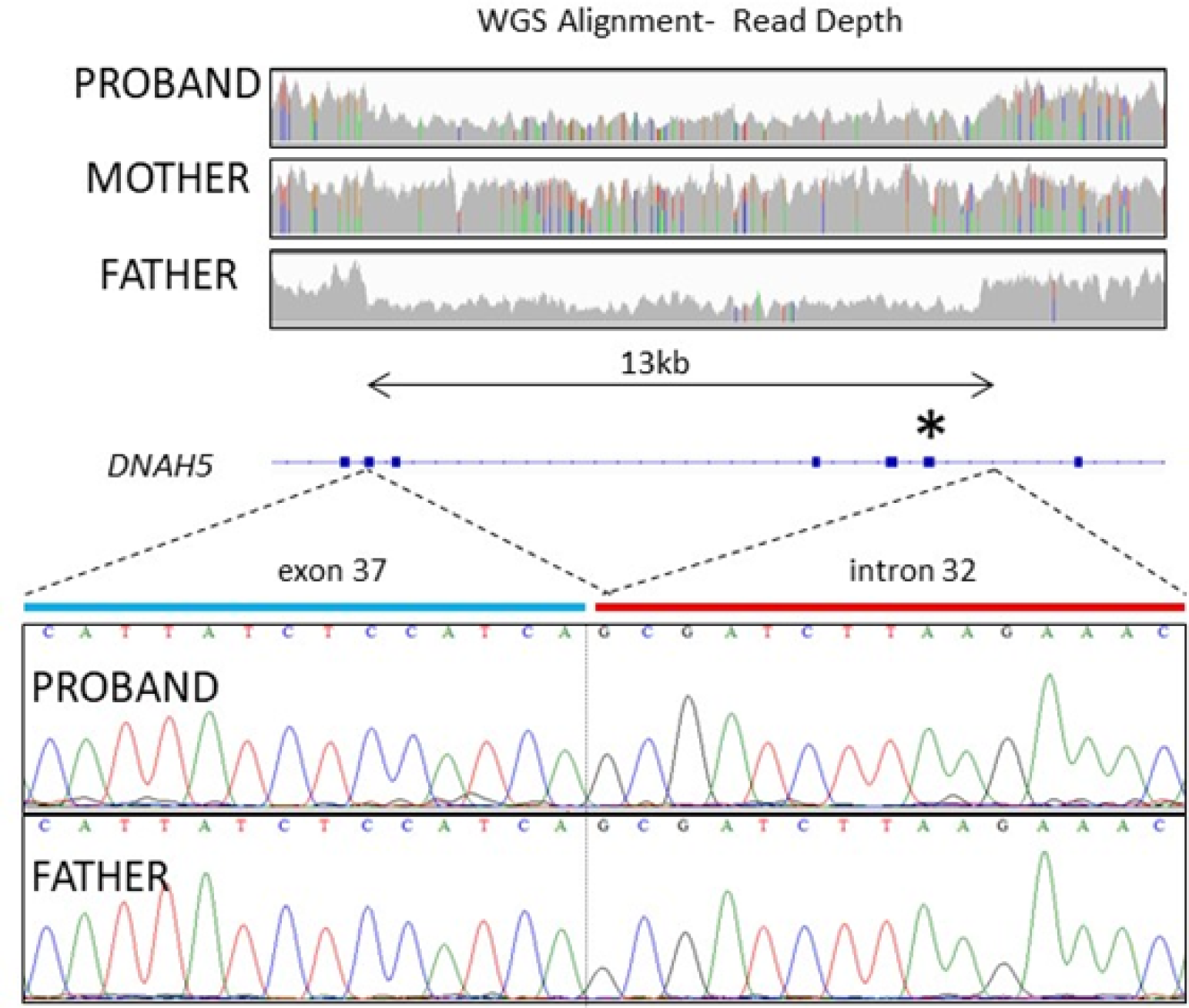
Sanger sequencing confirms 13kb deletion in *DNAH5* in Case 1. Alignments of the WGS data for Family 1 show a drop in read depth to approximately 50% of that of the surrounding regions across a 13kb region of *DNAH5* in the proband and the father. This spans from intron 32 to exon 37. PCR and Sanger sequencing across the breakpoints confirms this deletion. * indicates the position of the c.5281C>T nonsense variant, which is on the maternal haplotype and is therefore hemizygous in the proband.

**Supplementary Figure 4:**
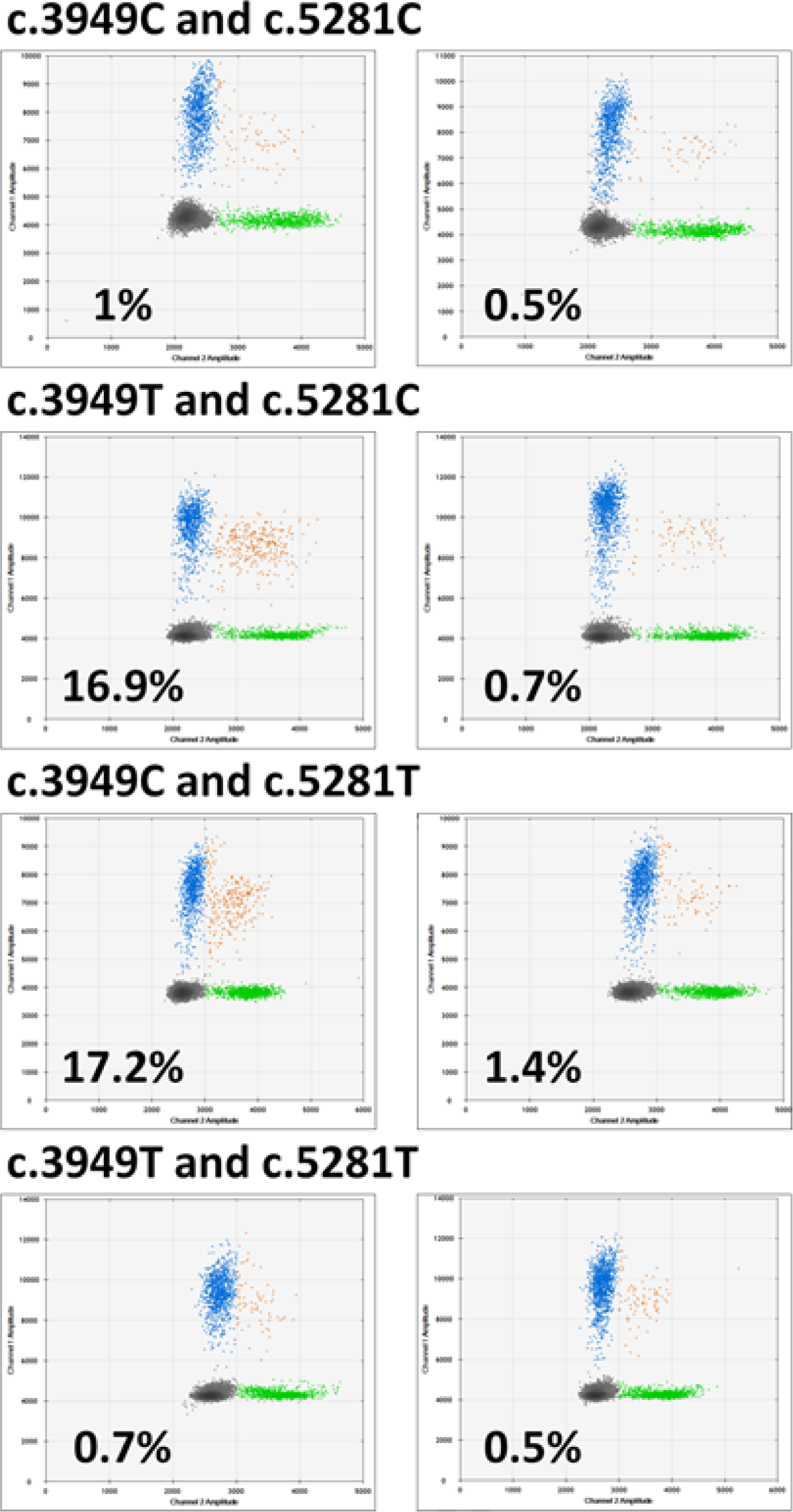
Drop phase results confirms the two *DNAH5* nonsense variants are on different haplotypes for Case 2. The c.3949 variant was assayed using FAM probes and the c.5281 variant was assayed using HEX probes. For each combination of alleles, a representative result from genomic DNA (left) and *Pac*I-digested DNA (right) is shown. Each figure plots the number of FAM-only positive (blue), HEX-only positive (green), FAM and HEX-positive (orange) and negative (grey) droplets. The linkage % is shown for each test. (Details see Supplementary File 3).

## SUPPLEMENTARY METHODS

### Patient cohort

Eight young people with PCD (50% female), aged 6 to 31 years (mean=15, SD= 7.9), were recruited to the study within the Department of Paediatric Respiratory and Sleep Medicine at the Royal Hospital for Sick Children, Edinburgh, and the South East Scotland Genetics service. All eight cases had a confirmed clinical diagnosis of PCD, following TEM and/or HSVM of a nasal brush biopsy sample. The screening and diagnostic testing was performed according to the PCD National Service protocols, with investigations including nNO, nasal brush biopsies analysed by HSVM for ciliary beat frequency and pattern and quantitative electron microscopy for ciliary ultrastructure. Clinical phenotypes are shown in **Table 1**. Blood samples were collected in EDTA tubes from the patients and parents, where available. DNA was extracted using the Chemagic DNA blood kit (Chemagen) or the Nucleon Bacc3 kit (GE Healthcare). Sample ethnicity was assessed using Peddy (v4.0.6) (1) (**Supplementary Figure 1**).

### Gene Panel, sequence data analysis and variant classification

BCBio-Nextgen (0.9.7) was used for alignment and variant detection. This used bwa mem (0.7.13) to align reads to the hg38/GRCh38 reference genome (2), samblaster (0.1.22) to mark duplicate fragments (3) and GATK (3.4-0-g7e26428) for indel realignment and base recalibration (4). GATK HaplotypeCaller was used to calculate genotype likelihoods. Joint genotyping and quality control, including kinship estimates to confirm sample relatedness, were performed using in-house pipelines with GATK (4.0.2.1) following the GATK best practices. Variants were annotated using Ensembl variant effect predictor (VEP 90) (5).

A bespoke gene panel of 146 genes was created (**Supplementary Files 1 and 2**), based on the PCD PanelApp panel (v1.14) and five additional genes identified in the literature (*CFAP300*, *DNAH6*, *DNAJB13*, *STK36* and *TTC25*) (6–8). Variants (SNPs and small indels) were filtered to retain only those within genes on the gene panel and then further filtered to identify candidate variants by inheritance model (both biallelic and monoallelic), transcript consequence, and population allele frequency using the G2P plugin for VEP (9). Variants were assessed using Alamut (v2.13) (10) and classified using the ACMG variant interpretation guidelines (11,12). Any variants classified as pathogenic or likely pathogenic were validated using Sanger sequencing and were submitted to ClinVar. Parental samples were used to determine the phase of compound heterozygous variants, except for Case 2, which used droplet digital PCR (ddPCR) for phasing (**Supplementary File 3**).

SVs were called using Manta (13) and Canvas (version 1.38) (14). The detection of SVs can be challenging and it is common practise to use complementary approaches to detect them (15). Manta detects SVs using discordant paired-end and split reads, whereas Canvas relies on changes in read coverage. We searched for any SVs present in our gene panel that were inherited from either parent. We also searched for de novo SVs for cases where both parents were available. SVs were confirmed *in silico* using SV-Plaudit (16), a tool for rapidly curating SV predictions, and/or using the Integrative Genomics Viewer tool (17). Candidate variants were confirmed in the laboratory by PCR and Sanger sequencing across the deletion breakpoints.

For Case 3, we did not find any diagnostic variants using our gene panel. As *FOXJ1* was only recently identified as a PCD gene and hence was not present on the panel, we searched for SNVs, indels and SVs in Case 3 in this gene, given the ciliary agenesis phenotype observed in this case is associated with *FOXJ1*. We also expanded our analysis for this case to a genome wide search for SNV and small indel candidates with Slivar (0.1.10), following the protocol for rare diseases (https://github.com/brentp/slivar/wiki/rare-disease Date accessed: January 2020). A small number of variants was detected (**Supplementary Table 2**) and none were identified that fitted the current modes of inheritance for PCD.

### Modelling of whole exome sequencing data

A whole exome sequencing (WES)-like subset of the WGS data was obtained by extracting only the reads mapping to the regions in the TWIST Exome Capture Kit (using samtools v1.6) from the BAM file for each sample. The capture region fully covers the exons affected by copy number variants (CNVs) identified based on the WGS data (**Supplementary Figure 2**). The WES CNV calling was performed using ExomeDepth (v 1.1.15) separately on each individual from the three families in which a pathogenic CNV was identified (Families 1, 5 and 6). As controls, we used WES-like subset data from the WGS data for the other samples in this project (total of 21), from which we excluded any members of the family currently being evaluated. As the WGS data was aligned to GRCh38, we generated a custom reference dataset (exons.GRCh38) required by ExomeDepth, to replace the dataset currently distributed with the ExomeDepth package (exons.hg19); exons.GRCh38 is based on the latest CCDS release (r 22) available for the GRCh38 human genome reference (Available at: www.ncbi.nlm.nih.gov/projects/CCDS/CcdsBrowse.cgi?REQUEST=SHOW_STATISTICS).

### Homology modelling of DNAH11 and location of missense variants

A homology model of the C-terminal region of the DNAH11 motor domain (residues 3348-4504) was built using PHYRE2 (18), based upon the cryo-electron microscopy structure of human cytoplasmic dynein-1 (PDB ID: 5NUG) (19). The effects of the mutations in the C-terminal domain (CTD) (residues 4124-4504) on protein stability were modelled with FoldX (20), using default parameters and calculated over 10 replicates. Sequences of human dynein genes were aligned with MUSCLE (21) and the sequence alignment was visualised with MView (22).

