## Supplementary File 3 for "High diagnostic rate of whole genome sequencing in primary ciliary dyskinesia"

**Supplementary File 3: Droplet digital PCR for variant phasing**

Case 2 was recruited to the study as a singleton and therefore parental samples could not be used to phase the two nonsense variants in *DNAH5*, which lie 26kb apart. The variants were phased using the ddPCR-based assay drop-phase [1]. This takes advantage of the emulsion PCR system of ddPCR, which ensures each droplet contains only a single target molecule, and the use of two different fluorescently-labelled allele-specific probes, to the two variants of interest, to determine the alleles at each variant position that are located on the same target molecule (i.e. are in cis). Alleles on the same haplotype should be on the same DNA molecule and occur in the same droplet, whereas alleles on different haplotypes should be on different DNA molecules and therefore occur in different droplets.

Primers pairs were designed to amplify the *DNAH5* c.3949C>T and c.5281C>T variants. FAM-labelled allele-specific probes were designed for c.3949C and c.3949T, whilst HEX-labelled probes were designed for c.5281C and c.5281T. Iowa Black quenched probes were used to reduce binding of the fluorescently labelled probes with the non-target allele. Each reaction contained: 1x ddPCR supermix for probes (no dUTP) (Bio-Rad); 1µM primers for the c.5281 locus; 0.9µM primers for the c.3949 locus; 330nM c.5281 probe; 250nM c.3949 probe; 1µM quencher probes; and 17ng DNA.

Droplets were generated using the QX200 automated droplet generator (Bio-Rad). The TX100 thermal cycler was used for PCR, before droplets were read on the QX200 droplet reader (Bio-Rad). QuantaSoft (v1.7.4) [2] was used to analyse the data and calculate the % linkage (i.e. the percentage of molecules showing linkage for the two alleles being tested).

Four assays were set up in triplicate, with each assay testing one of the four possible allele combinations. As an additional control, *Pac*I-digested genomic DNA from the proband was tested with all four allele combinations in triplicate. *Pac*I cuts the DNA between the c.3949 and c.5281 variants, physically separating the alleles onto separate target molecules and therefore into separate droplets.

As shown in **Supplementary Figure 4**, the two nonsense variant are on different haplotypes. For the c.3949C/c.5281C and c.3949T/c.5281T variant combinations, the number of double-positive droplets is low, highlighted by the lack of linkage (<1%). However, for the c.3949C/c.5281T and c.3949T/c.5281C combinations, the number of double-positive droplets is higher and the linkage is approximately 17%. This linkage is lost when *Pac*I-digested DNA is used as the input. This indicates that the two nonsense variants, c.3949T and c.5281T, are in trans and therefore there is biallelic loss of *DNAH5* for Case 2.

2.	Bio-Rad Laboratories, *QuantaSoft*. 2014.
